# Quantitative susceptibility mapping improves detection of striatal tau-PET signal in Aβ+ mild cognitive impairment

**DOI:** 10.1101/2022.08.16.22278811

**Authors:** Jason Langley, Murphy Shao, Ilana J. Bennett, Xiaoping P. Hu, the Alzheimer’s Disease Neuroimaging Initiative

**Affiliations:** Center for Advanced Neuroimaging, University of California Riverside, Riverside, CA, USA; Department of Bioengineering, University of California Riverside, Riverside, CA, USA; Department of Psychology, University of California Riverside, Riverside, CA, USA

**Keywords:** Mild Cognitive Impairment, Alzheimer’s Disease, ^18^F-AV1451 PET, quantitative susceptibility mapping

## Abstract

**Background:** The presence of β-amyloid (Aβ) extracellular plaques and hyper-phosphorylated tau neurofibrillary tangles characterize the pathology of Alzheimer’s disease (AD). The ^18^F-AV1451 radioligand allows for assessment of tau burden in vivo using positron emission tomography (PET). However, this radioligand also binds with iron and this off-target binding may obscure tau deposition in iron rich gray matter structures in the striatum. Here we tested whether, when controlling for iron, striatal tau burden is higher in individuals at increased risk of developing AD (i.e., with mild cognitive impairment and high amyloid burden, Aβ+ MCI) relative to cognitively normal older adults.

**Methods:** In 38 Aβ+ MCI and 41 biomarker negative control participants (Aβ-, APOE ε4 non-carrier) from ADNI, we analyzed magnetic resonance imaging (MRI) measures sensitive to iron (quantitative susceptibility mapping) and PET measures of the ^18^F-AV1451 radioligand binding sensitive to tau burden in high (bilateral putamen, globus pallidus, caudate nucleus) and low (thalamus, motor cortex) iron-containing gray matter regions.

**Results:** After controlling for tissue susceptibility, higher tau burden was found in the putamen and globus pallidus of Aβ+ MCI compared to the control group. Higher tau burden in the caudate nucleus of the Aβ+ MCI group was correlated with higher Alzheimer’s Disease Assessment Scale (ADAS) scores.

**Conclusions:** Controlling for iron allows for the assessment of tau burden in iron rich deep gray matter structures. Our findings suggest that Aβ burden increases the risk of developing AD-related tau pathology in the striatum and cognitive impairment.

## 1. Introduction

Alzheimer’s disease (AD) pathology is characterized by the presence of β-amyloid (Aβ) extracellular plaques and neurofibrillary tangles (NFTs) comprised of hyperphosphorylated tau.^1^ Tau deposition can be assessed *in vivo* using the radioligand ^18^F-AV1451 (tau-PET). However, this radioligand is known to bind to iron in addition to tau NFTs,^2^ with tau-PET uptake being positively associated with MRI-based measures sensitive to iron.^3-6^ Postmortem studies have reported the presence of NFTs in the striatum,^7,8^ but iron-related off-target binding may confound measurement of tau burden in these iron-rich deep gray matter structures (putamen, globus pallidus, caudate nucleus).

Mild cognitive impairment (MCI) is characterized by declines in cognitive performance that do not meet the threshold for dementia and those with elevated Aβ levels are at an increased risk of progression to AD. *In vivo* examination of striatal NFTs is of particular interest in this population since higher tau burden is associated with higher Aβ burden in these patients.^9,10^ However, this is challenging because of high iron levels in the striatum and off-target binding of the radioligand. Thus, accounting for off-target binding to iron in the striatum, measured using quantitative susceptibility mapping (QSM), may enable assessment of striatal tau burden in Aβ+ MCI relative to control participants as well as its relations to cognitive impairment, as seen with striatal Aβ burden.^11-13^

## 2. Methods

### 2.1 ADNI overview

Data used in this study were obtained from the ADNI database (adni.loni.usc.edu). The ADNI was launched in 2003 as a public-private partnership, led by Principal Investigator Michael W. Weiner, MD. Up-to-date information can be found at www.adni-info.org. The ADNI study was approved by the local Institutional Review Boards of all participating sites. Study subjects and, if applicable, their legal representatives, gave written informed consent at the time of enrollment for imaging data, genetic sample collection and clinical questionnaires.

### 2.2 Participants

The ADNI3 database was queried for individuals with tau-sensitive PET (^18^F-AV1451), T_1_-weighted, and multi-echo gradient echo (2D-GRE) MRI images at the same scanning visit, as well as Aβ and apolipoprotein E-ε4 (APOE ε4) allele status. From this cohort, we selected all individuals with a diagnostic status of MCI or control at the time of the visit, which included 38 Aβ positive (Aβ+) MCI participants and 41 control participants who were both Aβ negative (Aβ-) and had no APOE ε4 alleles (APOE ε4 non-carrier). Alzheimer’s Disease Assessment Scale cog-13 (ADAS13), mini-mental state examination (MMSE), and Montreal Cognitive Assessment (MOCA) scores were downloaded for each subject. Data were downloaded between December 2019 and January 2024.

### 2.3 MRI acquisition

Additional details regarding image acquisition parameters can be found at www.adni-info.org/methods. Briefly, all MRI data used in this study were acquired on Siemens Prisma or Prisma fit scanners. Anatomic images were acquired with an MP-RAGE sequence (echo time (TE)/repetition time (TR)/inversion time=2.98/2300/900 ms, flip angle=9°, voxel size=1.0×1.0×1.0 mm^3^, and GRAPPA acceleration factor=2) and were used for registration to common space and correction of partial volume effects in the PET data.

Iron-sensitive data were collected with a three-echo 2D gradient recalled echo (GRE) sequence (TE_1_/∆TE/TR = 6/7/650 ms, flip angle=20°, field of view=220×220 mm^2^, matrix size of 256×256, 44 slices, slice thickness=4.0 mm) and used for measurement of brain iron. Each participant’s scan parameters were checked in the dicom header and no deviations in scan parameters were found in the participants used in this analysis.

### 2.4 QSM processing

Susceptibility images were constructed from the 2D-GRE images. A brain mask was derived from the first echo of the magnitude data. Background phase was removed with harmonic phase removal using the Laplacian operator (iHARPERELLA).^14^ Susceptibility maps were then derived using an improved least-squares (iLSQR) method and Laplace filtering (truncation threshold of 0.04). All susceptibility images were processed in MATLAB (The MathWorks, Inc., Natick, MA, USA). Resulting susceptibility maps were aligned to each subject’s T_1_-weighted image using a rigid body transform derived from the magnitude image from the first echo. Susceptibility values were normalized to median susceptibility in the lateral ventricles.

### 2.5 PET acquisition and processing

The radiochemical synthesis of ^18^F-AV1451 was overseen by Avid Radiopharmaceuticals and distributed to qualifying ADNI sites, where PET imaging was performed according to standardized protocols. Participants received an injection of 10 mCi of tracer followed by 75 min uptake period outside of the scanner. Emission data were collected as 6×5min frames. PET with computed tomography imaging (PET/CT) scanners acquired a CT scan for attenuation correction; PET-only scanners performed a transmission scan following the emission scan.

PET imaging data were processed using the fMRIB software library (FSL) and PET partial volume correction (PETPVC) toolbox.^15^ Tau-PET scans were motion corrected, averaged, and then registered to the T_1_-weighted image using a rigid body transform with trilinear interpolation.^4^ Grey matter, white matter, and CSF maps were segmented from T_1_-weighted MRI images and used for partial volume correction. A combination of Labbé and region-based voxel-wise correction applied to reduce sensitivity to point spread function mismatch.^15^ The median standardized uptake value ratio (SUVR) was calculated using a reference in the inferior bilateral cerebellar cortex, avoiding the cerebellar vermis to minimize potential off-target AV1451 binding.^16^

### 2.6 Regions of interest

Regions of interest (ROIs) in bilateral putamen, globus pallidus, caudate nucleus, and thalamus were defined using the Harvard-Oxford subcortical atlas. A motor cortex ROI was defined using a previously published cortical atlas.^17^ ROIs were transformed from Montreal Neurological Institute (MNI) space to subject space using linear and nonlinear transforms in FSL as described in the earlier work^18^. Once in subject space, each ROI was thresholded at 60% and binarized. Mean susceptibility and tau-PET SUVR were measured in each resultant ROI for each participant.

### 2.7 Statistical analysis

All statistical analyses were performed using IBM SPSS Statistics software version 29 (IBM Corporation, Somers, NY, USA). A *P* value of 0.05 was considered significant for all statistical tests performed in this work. Normality of tau-PET and iron data was assessed using the Shapiro-Wilk test for each group and all data was found to be normal. The relationship between tau-PET SUVR and iron in striatal ROIs was assessed by Pearson correlations in each ROI. The effect of group (Aβ+ MCI, control) was tested with separate analysis of covariance (ANCOVA) in each striatal ROI (putamen, globus pallidus, caudate nucleus) for tau-PET SUVR, controlling for age, sex, site, education, and mean susceptibility in each ROI. As striatal Aβ burden is associated with AD-related cognitive decline,^11-13^ we elected to examine the effect of striatal tau burden on general cognition by correlating striatal tau-PET SUVR with general cognitive measures, controlling for susceptibility.

## 3. Results

### 3.1 Sample demographics

Demographic data for the control and Aβ+ MCI groups are shown in **Table 1**. The Aβ+ MCI group was significantly older and had worse performance across all cognitive measures than the control group (*Ps*<10^−3^), but there was no group effect for education (*P*=0.126). A significant difference in sex (χ^2^=4.904; *P*=0.027) and APOE ε4 carrier status (χ^2^=10.736; *P*=0.001) distribution was found between the Aβ+ MCI and control groups.

**Table 1.** Demographic and clinical characteristics of the control and Aβ+ MCI groups. Data are presented as mean ± standard deviation. Two-tailed *t*-tests were used for group comparisons of age, education, and cognition. Bold *t* values indicate significant comparisons. MoCA = Montreal cognitive assessment; MMSE = Mini-Mental State Exam. CDR = clinical dementia rating, ADAS = Alzheimer’s Disease Assessment Scale, APOE = Apolipoprotein E ε4 allele

| | Control | Aβ+ MCI | *t* | *P* |
| --- | --- | --- | --- | --- |
| N (M/F) | 41 (13/28) | 38 (22/16) | **5.481** | **0.019** |
| Age | 68.1 ± 9.0 | 74.8 ± 7.8 | **12.588** | **<10<sup>-3</sup>** |
| APOE ε4 carriers | 0 | 26 | **10.736** | **0.001** |
| Education | 15.9 ± 2.0 | 15.4 ± 2.7 | 0.912 | 0.343 |
| CDR Global | 0.0 ± 0.0 | 0.5 ± 0.2 | **356.276** | **<10<sup>-3</sup>** |
| MOCA | 25.5 ± 2.4 | 21.1 ± 3.6 | **40.346** | **<10<sup>-3</sup>** |
| MMSE | 29.0 ± 1.2 | 26.3 ± 2.4 | **40.485** | **<10<sup>-3</sup>** |
| ADAS Delayed Recall | 2.6 ± 1.7 | 6.3 ± 2.5 | **57.987** | **<10<sup>-3</sup>** |
| ADAS13 | 8.3 ± 4.0 | 17.6 ± 6.5 | **53.519** | **<10<sup>-3</sup>** |

### 3.2 Relationships with age

Age was not significantly correlated with susceptibility in the globus pallidus, caudate nucleus, thalamus, or motor cortex in any group (*P*s>0.322). However, age was significantly associated with putamen susceptibility in the control (*r*=0.299, *P*=0.025) group, but not in the Aβ+ MCI group (*r*=0.106, *P*=0.528).

Age was not significantly correlated with tau-PET SUVR in any region (*Ps*>0.074), and controlling for susceptibility did not significantly alter these correlations (*Ps*>0.100)

### 3.3 Tau-PET and susceptibility

Mean susceptibility and tau-PET SUVR images are shown in **Figure 1**. Significant correlations between striatal tau-PET SUVR and susceptibility were observed within both groups in the caudate nucleus (control: *r*=0.327, *P*=0.026; Aβ+ MCI: *r*=0.473, *P*=0.005), globus pallidus (control: *r*=0.470, *P<*10^−3^; Aβ+ MCI: *r*=0.424, *P*=0.013), and putamen (control: *r*=0.534, *P<*10^−3^; Aβ+ MCI: *r*=0.333, *P*=0.027). Correlations for the control and Aβ+ MCI groups are shown in Figure 2. Controlling for age did not substantially alter these associations in the Aβ+ MCI (*rs*>0.383; *Ps*<0.021) and control (*rs*>0.309; *Ps*<0.030) groups. Correlations between striatal tau-PET SUVR and susceptibility are shown **Figure 2**.

**Figure 1.**
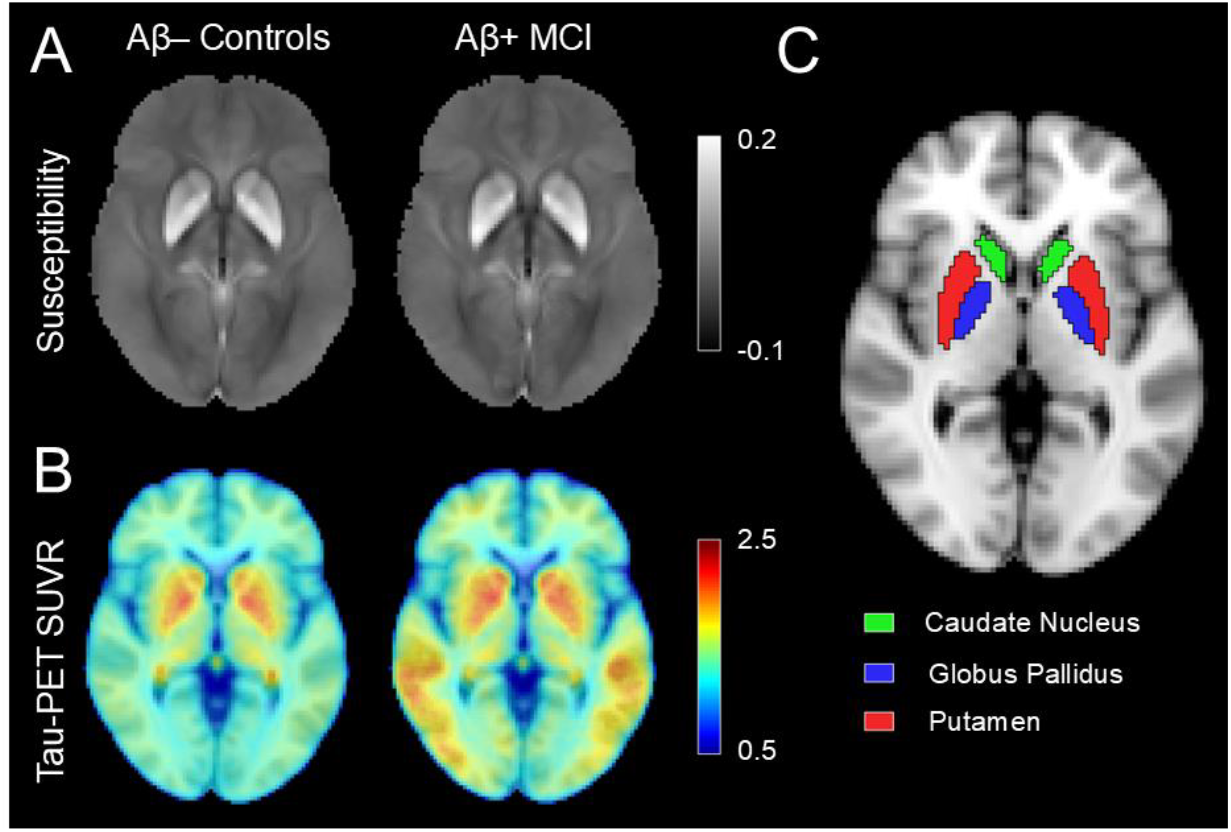
Mean tau-pet SUVR and mean susceptibility images at the level of the striatum in control and Aβ+ MCI groups are displayed in (A) and (B), respectively. ROIs for the caudate nucleus, globus pallidus, and putamen are colored in green, blue, and red, respectively, are shown in (C).

**Figure 2.**
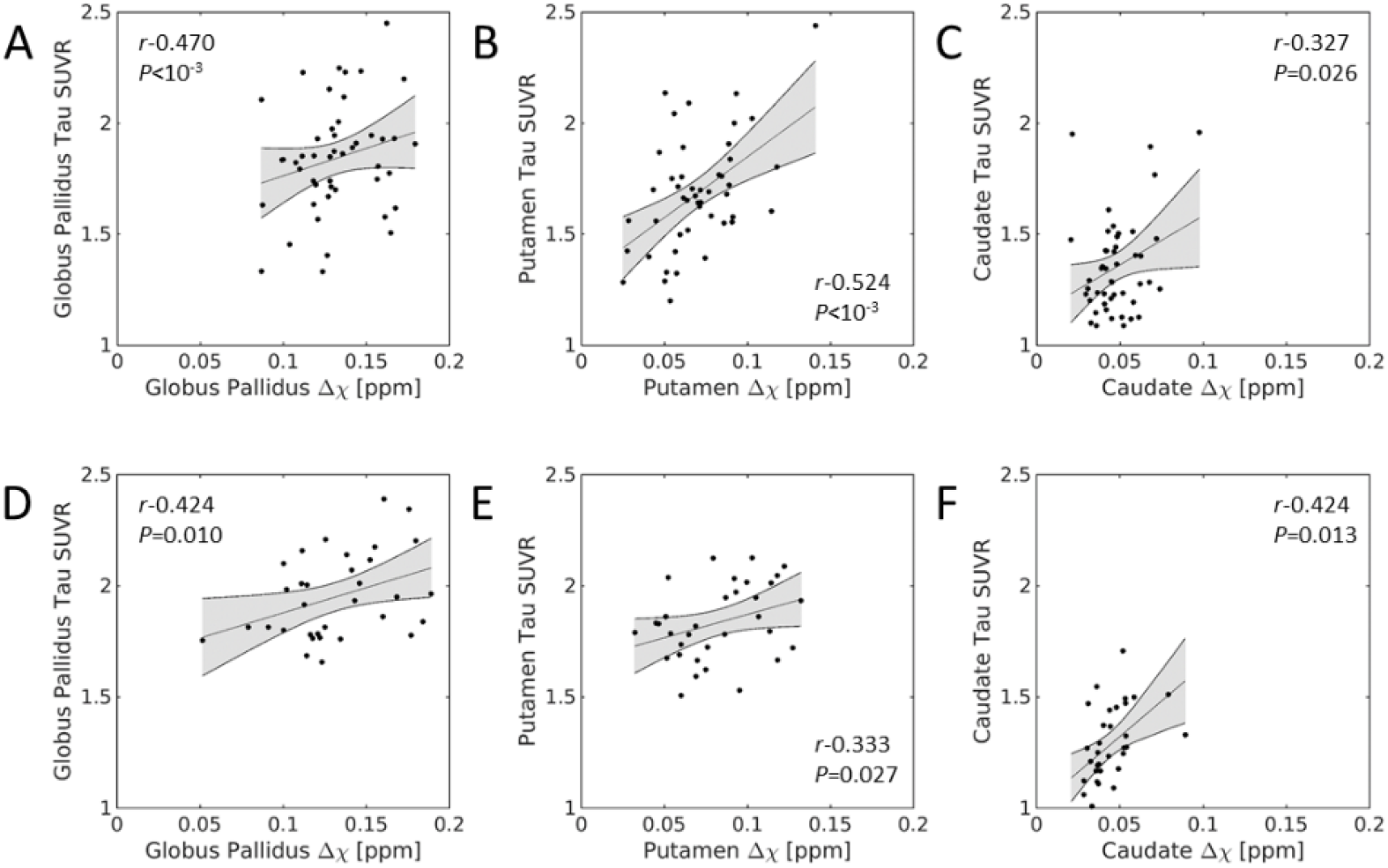
Correlations between susceptibility and tau-PET SUVR in the striatum are shown for the control group (A-C; top row) and Aβ+ MCI group (D-F; bottom row).

A significant negative correlation was seen between susceptibility and tau-PET SUVR in the thalamus of the control group (*r*=-0.376; *P*=0.004), but not in the Aβ+ MCI group (*r*=-0.308; *P*=0.060). No association was seen between susceptibility and tau-PET SUVR in the motor cortex of either group (*rs<*0.108; *Ps*>0.430).

### 3.4 Group differences in tau and susceptibility

Separate ANCOVAs were performed to examine the influence of off-target binding on striatal tau-PET SUVR, controlling for age, education, sex, and site, with and without inclusion of susceptibility as a covariate. A comparison of residualized tau-PET SUVR in the putamen, globus pallidus, and caudate nucleus is shown in **Figure 3**. In the models including susceptibility as a covariate, a significant main effect in group was observed with increases in tau-PET SUVR in the putamen (*F*=5.587; *P*=0.021) and globus pallidus (*F=*4.473; *P*=0.038), but not in the caudate nucleus (*F*=0.084; *P*=0.773), of the Aβ+ MCI group relative to the control group. Susceptibility (*Ps*<10^−3^) and sex (*Ps*<0.013) were significant covariates in the putamen and globus pallidus models.

**Figure 3.**
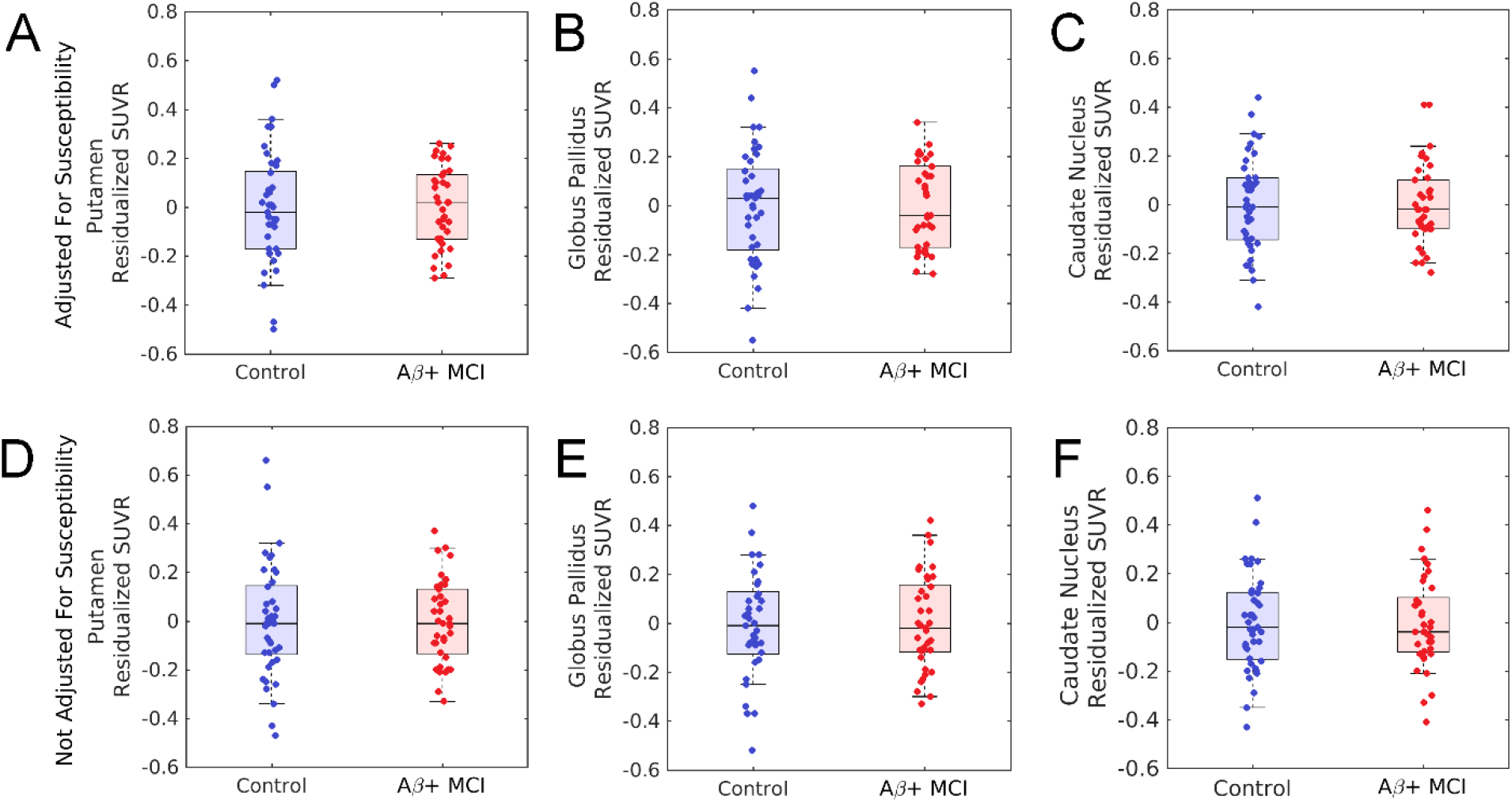
Boxplots of residualized tau-PET SUVR in the putamen, globus pallidus, and caudate nucleus for Aβ− control and Aβ+ MCI participants. SUVR values were residualized using linear models adjusting for age, sex, education, and site (bottom row; D-F), and additionally susceptibility (top row; A-C). Inclusion of susceptibility as a covariate increases separation between groups in the putamen and globus pallidus, whereas no group differences are observed in the caudate nucleus.

In the models excluding susceptibility as a covariate, a significant main effect in group was observed in the putamen (*F*=5.502; *P*=0.022) but not in the globus pallidus (*F*=2.083; *P*=0.153) or caudate nucleus (*F*=0.573; *P*=0. 451). In the putamen, sex (*P*=0.024) was a significant covariate with the Aβ+ MCI group showing elevated tau-PET SUVR relative to the control group. No group differences were seen in the thalamus or motor cortex for models with and without inclusion of susceptibility as a covariate (*Ps*>0.184).

Analyses repeated without partial volume correction yielded a similar pattern of results. Specifically, inclusion of susceptibility as a covariate remained associated with elevated tau-PET SUVR in the putamen (*F*=4.322; *P*=0.041) and globus pallidus (*F*=4.764; *P*=0.032) in the Aβ+ MCI group, while exclusion of susceptibility attenuated these effects, with group differences observed only in the putamen (*F*=4.984; *P*=0.029). No group differences were observed in the caudate nucleus (*Fs*<3.299; *Ps*>0.073). These findings indicate that partial volume correction did not materially influence the reported effects.

### 3.4 Effects of APOE ε4 in Aβ+ MCI

Demographic data for APOE ε4 carriers and non-carriers in the Aβ+ MCI is shown in **Table 2**. No difference in age, sex, education or cognition was observed between APOE ε4 carriers and non-carriers (*Ps*>0.437).

**Table 2.** Demographic information of the APOE ε4 carrier and non-carriers in the Aβ+ MCI group. Data are presented as mean ± standard deviation. Two-tailed *t*-tests were used for group comparisons of age, education, and cognition. Bold *t* values indicate significant comparisons.

| | Control | Aβ+ MCI | *t* | *P* |
| --- | --- | --- | --- | --- |
| N (M/F) | 41 (13/28) | 38 (22/16) | **5.481** | **0.019** |
| Age | 68.1 ± 9.0 | 74.8 ± 7.8 | **12.588** | **<10<sup>-3</sup>** |
| APOE ε4 carriers | 0 | 26 | **10.736** | **0.001** |
| Education | 15.9 ± 2.0 | 15.4 ± 2.7 | 0.912 | 0.343 |
| CDR Global | 0.0 ± 0.0 | 0.5 ± 0.2 | **356.276** | **<10<sup>-3</sup>** |
| MOCA | 25.5 ± 2.4 | 21.1 ± 3.6 | **40.346** | **<10<sup>-3</sup>** |
| MMSE | 29.0 ± 1.2 | 26.3 ± 2.4 | **40.485** | **<10<sup>-3</sup>** |
| ADAS Delayed Recall | 2.6 ± 1.7 | 6.3 ± 2.5 | **57.987** | **<10<sup>-3</sup>** |
| ADAS13 | 8.3 ± 4.0 | 17.6 ± 6.5 | **53.519** | **<10<sup>-3</sup>** |

In models excluding susceptibility as a covariate, no significant main effects of APOE status were observed for the putamen (*F*=2.914; *P*=0.098), globus pallidus (*F*=1.747; *P*=0.197), caudate nucleus (*F*=2.232; *P*=0.144), or motor cortex (*F*=2.537; *P*=0.122). Age, sex, site, and education (*Ps>*0.075) were not significant covariates in the model for any ROI. A significant main effect of APOE status was observed in the thalamus (*F*=4.519; *P*=0.042) with ε4 carriers showing higher tau-PET SUVR than to non-carriers. Age, sex, site, and education (*P=*0.415) were not significant covariates in the model.

When susceptibility was included as a covariate, significant main effects of APOE status were observed in the putamen (*F*=4.842; *P*=0.036) and caudate nucleus (*F*=4.519; *P*=0.046) but not in the globus pallidus (*F*=3.061; *P*=0.090). Susceptibility was a significant covariate in each model (*Ps*<0.019) whereas age, sex, site, and education (*Ps*>0.066) were not. No significant main effect was seen for the thalamus or motor cortex when susceptibility is included in the model (*Ps*>0.172).

### 3.4 Relationships to Cognition

Significant correlations between putamen tau-PET SUVR and ADAS13 were observed in both the control (*r*=0.323; *P=*0.029) and Aβ+ MCI (*r*=0.304; *P=*0.048) groups, controlling for age, sex, site, education and susceptibility. A significant correlation was observed between caudate tau-PET SUVR and ADAS13 in the control group (*r*=0.284; *P=*0.049) but not in the Aβ+ MCI group (*r*=-0.112; *P=*0.275), controlling for age, sex, education, and susceptibility. No significant correlations were observed between MOCA or MMSE score and tau-PET SUVR in any striatal ROI for either group (*Ps*>0.082).

## 3. Discussion

The ^18^F-AV1451 radioligand binds to iron in addition to tau neurofibrillary tangles.^2^ This off-target binding can contribute to radioligand uptake and may confound assessment of tau burden in iron-rich subcortical structures. In this study, we used susceptibility, an MRI measure sensitive to iron content,^19^ to assess and control for off-target binding effects associated with the ^18^F-AV1451 radioligand in iron-rich gray matter nuclei. Significant positive correlations were observed between iron and tau-PET SUVR in striatal ROIs (globus pallidus, putamen, and caudate nucleus). In the thalamus, a significant negative correlation was observed in the control group. Elevated tau-PET SUVR was observed in the putamen and caudate nucleus of Aβ+ MCI participants relative to controls when susceptibility was included as a covariate, but not when susceptibility was excluded from the model. After controlling for susceptibility, elevated tau-PET SUVR was seen in the putamen and caudate nucleus of APOE ε4 carriers relative to non-carriers within the Aβ+ MCI group.

In iron-rich grey matter nuclei, higher iron content is reflected by higher susceptibility values.^19,20^ Binding of the ^18^F-AV-1451 radioligand has been observed in iron-rich grey matter nuclei^2,21^ and prior studies have reported positive correlations between susceptibility and tau-PET SUVRs in striatal ROIs of control and MCI participants.^5,6^ Consistent with these findings, we observed significant positive correlations between susceptibility and tau-PET SUVR in all striatal regions (globus pallidus, putamen, caudate nucleus) in both control and Aβ+ MCI groups. No such associations were observed in regions with lower iron content (i.e. motor cortex and thalamus), supporting the interpretation that iron contributes to tau-PET signal in iron-rich grey matter nuclei.

Postmortem studies have found neurofibrillary tangles in the striatum of patients in the advanced stages of Alzheimer’s disease.^7,8^ However, *in vivo* tau-PET studies have not reported robust differences in striatal tau burden, likely due to off-target binding of first-generation tau tracers, such as ^18^F-AV1451, in iron-rich structures.^22,23^ Consistent with this, we did not observe elevated tau burden in Aβ+ MCI participants relative to controls in models that did not account for susceptibility. In contrast, inclusion of susceptibility as a covariate revealed significant group differences in the putamen and globus pallidus. These findings suggest that accounting for iron-related off-target binding using quantitative susceptibility mapping enables the detection of biologically meaningful tau-related signal in the striatum, consistent with prior neuropathological observations.

Individuals carrying the APOE ε4 allele are at increased risk of developing AD pathology^24^ and ε4 carrier status is associated with greater tau accumulation.^25-27^ We observed elevated tau-PET SUVR in the putamen and caudate nucleus in APOE ε4 carriers relative to non-carriers in the Aβ+ MCI group, but only after controlling for susceptibility. This finding suggests that iron-related off-target binding may obscure genotype-related differences in tau deposition in iron-rich regions. Accounting for susceptibility revealed APOE-associated effects consistent with its established role in promoting AD pathology, indicating that correction for iron-related signal may improve detection of tau-related signal when using first-generation tau radioligands in subcortical structures.

Interestingly, caudate tau-PET SUVR was significantly correlated with ADAS13 scores in the Aβ+ MCI group, with higher tau burden associated with lower cognitive performance. While we did not directly assess Aβ burden in the striatum, prior studies have demonstrated that striatal Aβ deposition is associated with cognitive decline in AD.^11-13^ These findings support the relevance of striatal pathology to cognitive impairment and suggest that tau deposition in these regions may contribute to cognitive dysfunction. These results should be interpreted with caution given the complexity of cognitive measures and the relatively small sample size.

This study has several caveats. First, QSM is sensitive not only to iron, but also to other sources of magnetic susceptibility, including myelin and calcium.^19,28^. We focused on iron-rich grey matter regions (globus pallidus, putamen, caudate nucleus) and these nuclei are permeated by small white matter bundles, which may introduce partial volume effects and influence susceptibility measurements.^29-31^ Second, while iron accumulation has been implicated in promoting tau hyperphosphorylation and aggregation,^32-34^ off-target binding of the ^18^F-AV1451 radioligand complicates interpretation of the relationship between susceptibility and tau aggregation. As such, disentangling the relationship between iron and tau using first generation tau tracers remains challenging.

Overall, these findings demonstrate that inclusion of susceptibility materially alters the interpretation of tau-PET signal in iron-rich subcortical structures. By accounting for iron-related off-target binding, we revealed Aβ group differences and APOE ε4–associated effects that were not detectable using conventional analyses. These results highlight the importance of integrating MRI-based susceptibility measures when studying tau deposition in deep gray matter using the ^18^F-AV-1451 radioligand. More broadly, this approach provides a framework for improving the specificity of molecular imaging signals in regions prone to off-target binding, with implications for both mechanistic studies and clinical applications in early AD.

## Data Availability

All data supporting these analyses are available through the Alzheimer's Disease Neuroimaging Initiative (ADNI).

## Acknowledgements

The authors would like to thank Sumanth Dara for assistance in organizing the data set. Data used in preparation of this article were obtained from the Alzheimer’s Disease Neuroimaging Initiative (ADNI) database (adni.loni.usc.edu). As such, the investigators within the ADNI contributed to the design and implementation of ADNI and/or provided data but did not participate in analysis or writing of this report. A complete listing of ADNI investigators can be found at: http://adni.loni.usc.edu/wp-content/uploads/how_to_apply/ADNI_Acknowledgement_List.pdf

